# Personal Involvement with Celiac Disease as a Motive for Research among Celiac Disease Researchers

**DOI:** 10.1101/2024.09.22.24314161

**Authors:** Marcela Banegas, Rebecca Sprague, Carolina Vigna, Jose Emmanuel González, David Flores-Marín, Cesar Palacios, Jocelyn Silvester, Amelie Therrien

## Abstract

**Introduction:** We investigated the frequency of personal involvement with celiac disease (CeD) among CeD researchers and their motive to pursue CeD research, compared to researchers in inflammatory bowel disease (IBD) or type 1 diabetes (T1D) research.

**Methods:** An online survey was sent to CeD, IBD and T1D researchers.

**Results:** 42% of CeD researchers had an involvement with their research interest, which was similar to T1D (51%) and significantly higher than the IBD respondents (27%, p<0.02).

**Discussion:** Personal involvement is common among the CeD researchers. Personal involvement with the disease may be beneficial when conducting patient-oriented research.

## INTRODUCTION

The motivation to go into a specific research field varies and may depend on several factors – including researcher’s interests, characteristics of the topic and type of research involved or random opportunity.

Chronic diseases may be linked to researchers at a personal level. Celiac disease (CeD) is a common, immune-mediated chronic condition affecting about 1% of the population and frequently diagnosed at an early age ^1,2^. Even though CeD has a huge unmet medical need with the gluten-free diet (GFD) being a high burden to the affected individuals, CeD research community is relatively small, with limited federal funding ^3^. We evaluated the frequency of personal involvement with CeD among the CeD research community and its role as a motive to go into CeD research. Additionally, we compared the degree of personal involvement to researchers in inflammatory bowel disease (IBD), another gastrointestinal immune condition, and type 1 diabetes (T1D), a non-gastrointestinal autoimmune disease that, similar to CeD, usually develops during childhood and adolescence.

## METHODS

Names and emails of the first three and last authors of PubMed-indexed articles published from July 1^st^ 2018 to June 30^th^ 2019 with the keyword “celiac disease”, “inflammatory bowel disease” and “type 1 diabetes” were retrieved using the easyPubMed package in R-studio ^4,5^. Emails were also collected manually using the Google search engine. Authors were excluded if they have only published (1) case-reports, (2) articles overlapping two or more research fields, (3) articles related to gluten/cereals without clear application in celiac disease (4) without any public contact information (email).

The online-platform Alchemer was used to conduct an online survey that included items related to motivations for a research career and personal involvement with the disease studied. Email invitations were sent to all included CeD, IBD and T1D researchers. The degree of personal involvement was defined as a personal diagnosis with the disease, having a family member with the disease and/or having someone in their research team with the disease. Chi-square statistical analysis was used to compare personal involvement among the three (CeD, IBD, T1D) research communities. This study was approved by the Institutional Review Board at Boston Children’s Hospital.

## RESULTS

A total of 2,948 email invitations were sent among the different research communities of which 315 researchers responded.

941 emails were sent to the CeD research community, 18% (173) were undeliverable (bounced emails or absent leaves); yielding a total of 768 delivered emails and 165/768 (21%) complete responses. A total of 2007 emails were sent to the IBD and T1D research communities, of which 22% were undeliverable (434/1086 IBD emails; 155/921 T1D emails). Complete responses were received from 75/807 (9.2%) IBD researchers and 75/766 (9.8%) TID researchers who were successfully contacted.

IBD and T1D researchers who responded were predominantly male [64% (48/75) IBD; 56% (42/75) T1D] while more CeD researchers were female [52% (86/165)] (See Table 1). Age distribution was similar across the three research fields, with many being 35-50 years old [49% (74/165) CeD; 53% (40/75) IBD; 44% (33/75) T1D. CeD respondents were commonly Junior Researcher/Associate Professor/Assistant (42%, 70/165) while IBD and T1D researchers were most commonly Professor/Senior researcher [44% (33/75) IBD, 55% (41) T1D]. T1D respondents were more often pediatric clinical researchers (32%, 24/75) while CeD and IBD respondents were adult clinical researchers [30% (49/165) CeD, 52% (39/75) IBD].

**Table 1.** Baseline Characteristics among research participants in Celiac Disease, Inflammatory Bowel Disease and Type 1 Diabetes research fields.

|  | Celiac<br>Disease<br>n (%) | Inflammatory<br>Bowel<br>Disease n (%) | Type 1<br>Diabetes<br>n (%) |
| --- | --- | --- | --- |
| **Sex** |  |  |  |
| Female | 86 (52) | 26(35) | 33(44) |
| Male | 77 (47) | 48(64) | 42(56) |
| Prefer not to answer | 2 (1) | 1 (1) |  |
| **Age group** |  |  |  |
| 18-34 | 23(14) | 6(8) | 3 (4) |
| 35-50 | 74(49) | 40(53) | 33(44) |
| 51-65 | 46(28) | 22(29) | 27(36) |
| >65 | 19(12) | 7(9) | 12(16) |
| Prefer not to Answer | 3 (2) |  |  |
| **Work Position** |  |  |  |
| Professor / Senior Researcher | 67 (41) | 33(44) | 41 (55) |
| Assistant/Associate professor / junior Researcher | 70 (42) | 30(40) | 26(35) |
| Graduate Student/ Post-doctoral fellow | 12 (7) | 5(7) | 4(5) |
| Other student (medical student, undergraduate, other) | 1(1) | 1(1) | 2(2) |
| Statistician | 1(1) | 1(1) | 2(2) |
| Other | 14 (8) | 5(7) | 2(3) |
| **Type of Research** |  |  |  |
| Basic Science Research | 27 (16) | 9(12) | 11(15) |
| Translational Research | 40(24) | 9(12) | 15(20) |
| Epidemiologic Research | 10 (6) | 2(3) | 10(13) |
| Clinical Research - Adults | 49(30) | 39(52) | 11(15) |
| Clinical Research - Children | 32 (19) | 14(19) | 24(32) |
| *Other | 7 (5) | 2(3) | 4(5) |

\*Other types of research: Plant/Breeding, Nutrition, Bioinformatics, Economics, Health Services

Overall, nearly 41% (128/315) of researchers reported having a degree of personal involvement with the disease. Fewer IBD researchers had a personal connection to the disease they were studying compared to CeD and T1D [27% (20/75) IBD; 42% (70/165) CeD; 51% (38/75) T1D]; p<0.02. A personal diagnosis of the disease studied was reported by 14% (10/70) in the CeD group, 5% (4/20) in the IBD group and 3% (2/38) in the T1D group; non statistically significant (p =0.245). Research team members were affected with the disease in 25% (41/165) of the CeD group, 39% (29/75) of the IBD group and 9% (7/75) of the T1D group. Some researchers reported having someone in their family with the diagnosis [11%(n=8) of the T1D, 5% (n=8) of the CeD and 7% (n=5) of the IBD researchers] (Figure 1).

**Figure 1.**
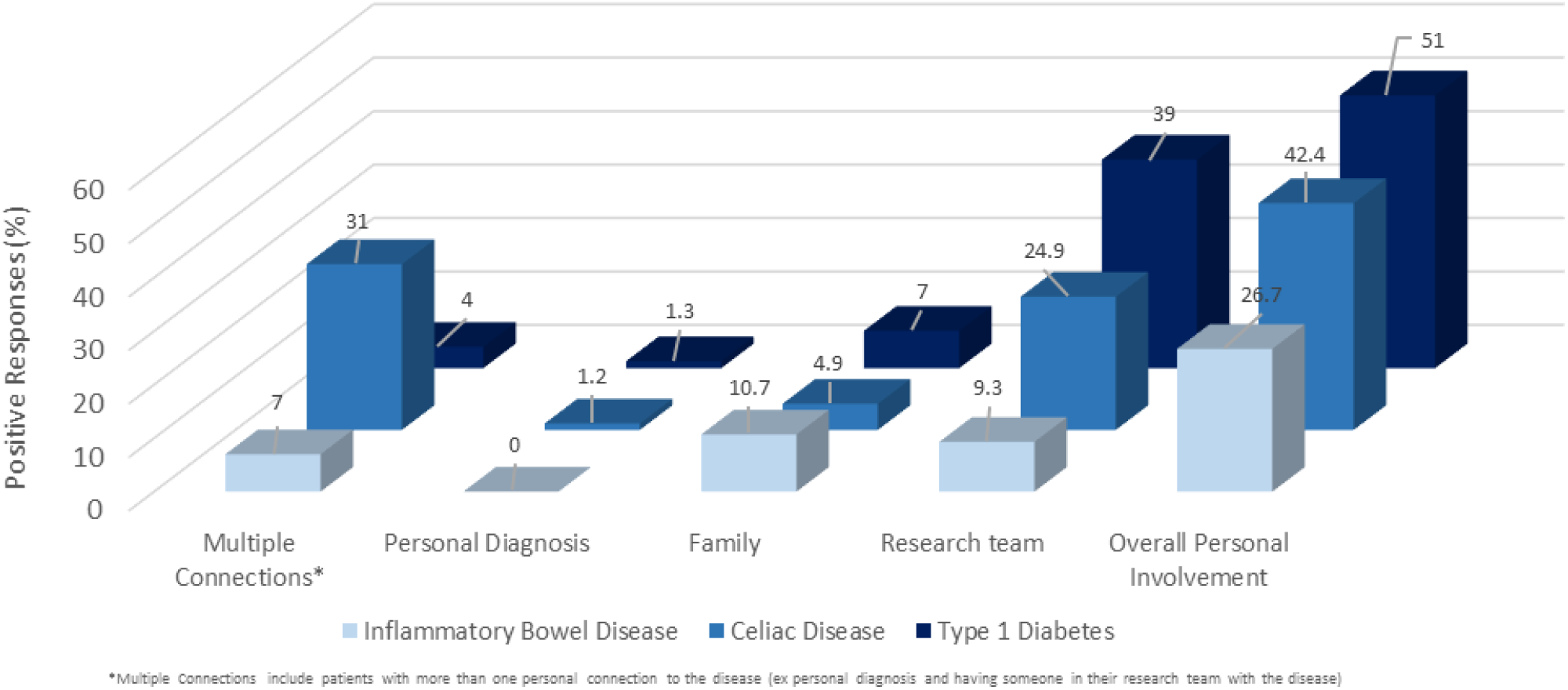
Degree of Personal Involvement among Celiac Disease, Inflammatory Bowel Disease and Type 1 Diabetes.

Finally, we asked the participants to rank their motives for going into the research field; “interest in disease pathophysiology” was ranked first by all three groups [33% (52/165) CeD, 37% (27/75) IBD and 37% (27/75) T1D] followed by “unmet medical need”, “career opportunity” and, finally, “personal involvement” (Figure 2).

**Figure 2.**
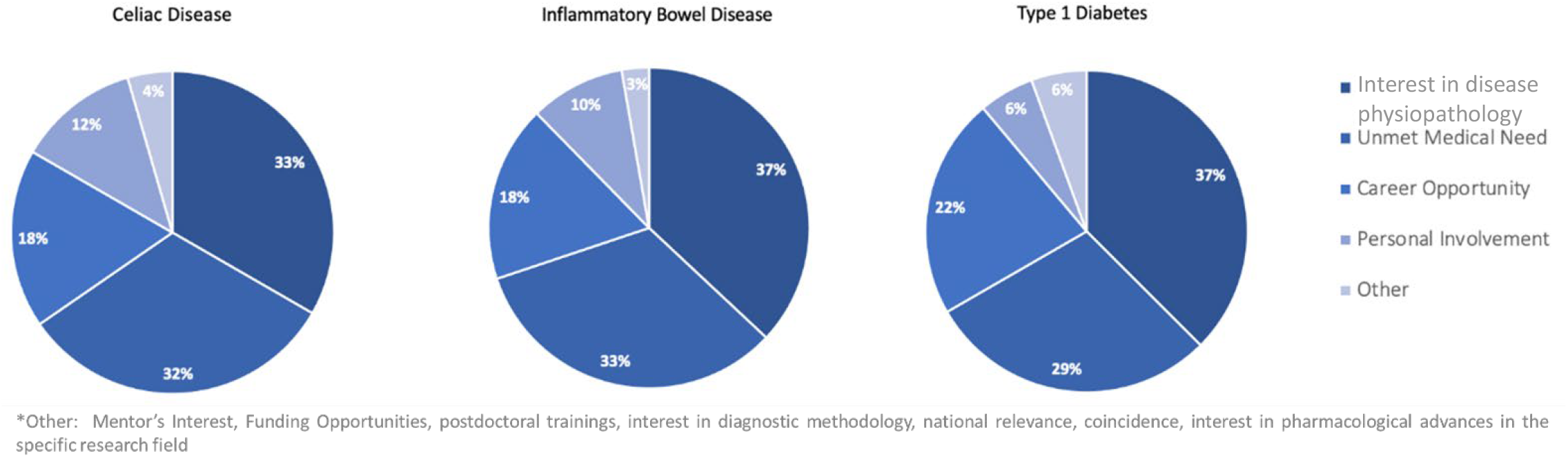
First Rank Motives of Going into the Research Field Among Celiac Disease, Inflammatory Bowel Disease and Type 1 Diabetes Researchers.

## DISCUSSION

We found that researchers commonly have personal involvement with the disease they study, particularly among the CeD (42%) and T1D (51%) research communities. Personal involvement was reported by fewer IBD researchers (27%) compared to the other groups; however, it remains prevalent. Researchers that reported having a personal connection might allow them to have a stronger patient perspective thus, influencing the orientation of their research however, this requires further investigation. Importantly, most researchers reported conducting clinical research which may allow them to conduct patient-centered research.

Academic training among researchers and their enthusiasm towards personal research interests might have an impact on the disagreement towards the relevancy of the researcher vs patient research perspective^6^. A 2014 publication in JAMA brings attention to this matter; stating that researchers and clinicians may not always be the best advocates to represent patient perspective ^7^. Having researchers with a personal connection to the disease they are studying allows them to incorporate both the research and patient perspective and further enhance research that may change on-going clinical practice. Nonetheless, having a personal connection and conducting research may also bring up ethical disadvantages^8^.

To our knowledge there are no prior studies reporting on the motivation factors of CeD researchers to go into the CeD research field. Even though this novel study provides results based on a diverse international cohort including responses from researchers from different ages, sex and type of research involved; it has its limitations. First, we were expecting a higher response rate from the research communities and were not expecting a difference in response rate across the three research groups. Second, risk of selection bias is possible due to the English-language restriction and the lack of accessibility to authors who don’t have their emails listed publicly. Finally, being a pioneer-study, the study design and the subjectivity of the outcome were hard to assess.

Personal involvement among CeD researchers is common among the CeD research field. The impact of personal involvement in the research being conducted needs further investigation. Even though personal involvement was not reported as the main motivation factor to go into the research field, personal involvement with the disease may be beneficial when conducting patient-oriented research.

## Data Availability

All data produced in the present study are available upon reasonable request to the authors

## Notes

**Disclosures:** The authors have no relevant disclosures or conflicts of interest.

### Competing Interest Statement

The authors have declared no competing interest.

### Funding Statement

Amelie Therrien was supported by FRQS Phase 2 Fellowship training Award, Douglas Kinnear Award, American College of Gastroenterology Career Development Award
Marcela Banegas, Carolina Vigna, David Flores, Jose Emmanuel Gonzalez, Cesar Palacios were supported by International Research Initiative
Jocelyn Silvester was supported by NIDDK 1K23DK119584-01

### Author Declarations

This study was approved by Boston Children's Hospital Institutional Review Board.

## REFERENCES

1. Lebwohl B, Sanders DS, Green PHR. Coeliac disease. The Lancet. 2018;391(10115):70–81. doi:10.1016/S0140-6736(17)31796-8

2. Singh P, Arora A, Strand TA, et al. Global Prevalence of Celiac Disease: Systematic Review and Meta-analysis. Clin Gastroenterol Hepatol. 2018;16(6):823-836.e2. doi:10.1016/j.cgh.2017.06.037

3. Clerx E, Kupfer SS, Leffler DA. Disparities Among Gastrointestinal Disorders in Research Funding From the National Institutes of Health. Gastroenterology. 2017;153(4):877–880. doi:10.1053/j.gastro.2017.08.051

4. Damiano, Fantini. EasyPubMed: Search and Retrieve Scientific Publication Records from PubMed.; 2019. https://CRAN.R-project.org/package=easyPubMed.

5. R Core Team. R: A Language and Environment for Statistical Computing. Vienna, Austria: R Foundation for Statistical Computing; 2021. https://www.R-project.org/.

6. Methodology Committee of the Patient-Centered Outcomes Research Institute (PCORI). Methodological standards and patient-centeredness in comparative effectiveness research: the PCORI perspective. JAMA. 2012;307(15):1636–1640. doi:10.1001/jama.2012.466

7. Frank L, Basch E, Selby JV, For the Patient-Centered Outcomes Research Institute. The PCORI Perspective on Patient-Centered Outcomes Research. JAMA. 2014;312(15):1513–1514. doi:10.1001/jama.2014.11100

8. Yanos PT, Ziedonis DM. The Patient-Oriented Clinician-Researcher: Advantages and Challenges of Being a Double Agent. Psychiatr Serv Wash DC. 2006;57(2):249–253. doi:10.1176/appi.ps.57.2.249

